# Prevalence of AI findings on Chest X-ray in patients with lung cancer: a cross-sectional cohort study

**DOI:** 10.1101/2024.12.20.24319410

**Authors:** Rhidian Bramley, Philip Broadhurst, Anna Sharman, Dennis Robert, Elodie Weber, Marcus Simon, Rehan Nasser, Louise Brown, Seamus Grudy, Matthew Evison

## Abstract

**Background:** Chest X-ray Radiography (CXR) is the primary investigation for patients with potential symptoms of lung cancer in the UK. Artificial intelligence (AI) can detect abnormalities on CXR and prioritise cases for reporting. We describe a method to determine which AI findings are associated with lung cancer to inform and validate prioritisation strategies.

**Methods:** This multicentre study compared the prevalence of AI findings on CXR in a retrospective cohort of patients diagnosed with lung cancer (4408 CXR) to the prevalence of AI findings in a prospective cohort of CXR from the referral population (107,065 CXR). Nineteen AI findings were assessed individually and in combination.

**Results:** The most common AI findings in patients with lung cancer compared to the referral population were ‘Abnormal’ (92.6% vs 60.9%), Opacity (83.4% vs 45.4%), Consolidation (36.9% vs 12.9%), Atelectasis (33.5% vs 20.9%) and Nodule (32.7% vs 10.9%). The finding most associated with cancer based on the prevalence ratio in the cancer and referral cohorts were ‘Lung nodule malignancy’ (13.3), Cavity (4.0), Tracheal deviation (3.1), Nodule (3.0) and Consolidation (2.9). The percentage of CXR classed as ‘AI Abnormal’ varied by the referral cohort, 63.5% from Accident & Emergency vs 43.0% from General Practice. This suggests significant variation in the complexity of cases across referral pathways.

**Conclusion:** Individual AI findings had limited sensitivity in detecting lung cancer. Using combinations of AI findings significantly improved cancer detection rates but required prioritising a larger proportion of CXR from the referral population.

**Key messages:** 

**What is already known on this topic:** The National Institute for Clinical Excellence (NICE) early value assessment of “Artificial intelligence-derived software to analyse CXR for suspected lung cancer in primary care referrals” has identified the need for more research on the ability of AI to identify normal and abnormal findings on CXR to enable prioritised reporting of and speed up the cancer pathway.

**What this study adds:** This study describes a method that sites deploying AI can use to determine the relative prevalence of AI finding on CXR in patients with lung cancer and in the referral population. The results indicate that prioritisation approaches focusing on single abnormalities such as lung nodule detection will miss most lung cancers on CXR.

**How this study might affect research, practice or policy:** Prioritisation strategies need to optimise the detection of lung cancer and other significant pathology. Using AI to prioritise abnormal CXR requires ethical consideration as moving some patients to the front of the reporting queue can also have a detrimental effect on patients that are not prioritised. This study indicates a range of AI findings need to be prioritised to maximise the detection rate of lung cancer.

## Introduction

Lung cancer is the third most common cancer diagnosed in England, but accounts for more deaths than other malignancies [1] . Chest X-ray (CXR) is the primary investigation for patients with symptoms of lung cancer in the UK [2]. Patients with findings suspicious for lung cancer on CXR are referred for CT scan assessment and biopsy to make the diagnosis [3].

The National Optimum Lung Cancer Pathway [NOLCP] recommends CXR should be reported immediately or within the first 24 hours from acquisition [4]. Data from NHS England shows only 24% of CXR are reported the same day [5]. Delays in the diagnostic pathway influence patient experience and can shorten life expectancy [6] [7]. Prioritisation of abnormal CXR from primary care for immediate reporting can significantly improve turnaround times and halve the time to diagnosis of lung cancer [8] [9]. However, there is limited evidence on how AI can be optimised for such prioritisation without unintended consequences for non-prioritised cases.

Artificial Intelligence (AI) can be used to prioritise patients with abnormal findings to the front of the reporting queue. The NICE early value assessment report on use of CXR AI for suspected lung cancer in primary care has recommended more research is needed on key outcomes including the diagnostic accuracy of AIIZlderived software and the impact of this on work prioritisation and patient flow [10].

To address this gap, our study sought to determine the prevalence of AI findings in patients with lung cancer and in a representative referral population, to help inform the strategy for prioritising reporting of CXR and the percentage of patients that would need to be prioritised.

## Methods

The study was conducted by the Greater Manchester Cancer Alliance as a part of a collaborative working agreement and service evaluation with Sectra the Picture Archive and Communication System (PACS) supplier, Qure.ai the AI software provider, and AstraZeneca the project sponsor.

The AI software (qXR version 3.1 from Qure.ai) was deployed via the Sectra amplifier platform and configured to process CXR from seven acute NHS hospital trusts in Greater Manchester (total population 2.8 million). Each CXR was processed on the platform and the output from the AI returned to the clinical PACS system and Secure Data Warehouse (SDW) for analysis. The AI software was licensed to process the frontal chest radiograph and provides a list of up to 18 individual AI findings and ‘Abnormal’ signifying the presence of any of the AI findings. The manufacturer defined pre-set operating points were used.

An initial prioritisation grouping of AI findings was proposed by the project clinical advisory group comprising specialist lung clinicians and radiologists, based on the clinical suspicion of cancer and requirement for prioritisation (Table 1). An “or” condition was used for prioritisation. For example, if any one of the five findings (lung nodule, hilar prominence, mediastinal widening, cavity, and lung nodule malignancy) in Priority 1 group were detected by AI in a CXR, then it is flagged as Priority 1. Priorities 1 to 4 were also assigned an ordered hierarchy such that Priority 1 is the highest in the hierarchy. For example, if multiple findings across Priority 1, 2 and 3 are detected by AI in a CXR, then that CXR was flagged as Priority 1 and not Priority 2 or 3. The study was designed so it can be performed in ‘shadow mode’ (running in background without impacting workflow) on real world data, enabling sites to model how AI will perform in its intended purpose prior to clinical deployment.

**Table 1:**
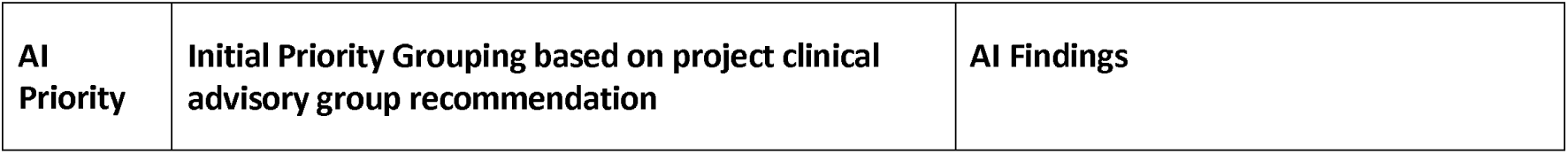
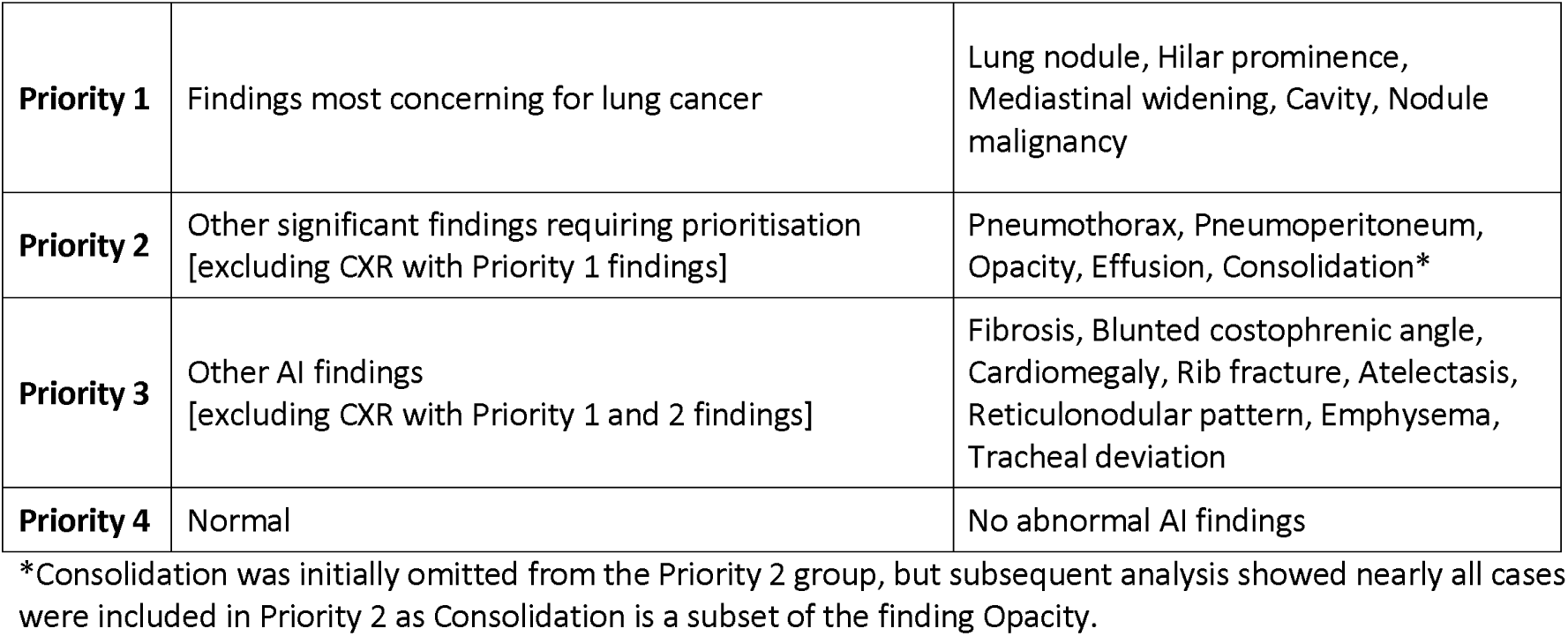
Initial priority grouping of AI findings recommended by the project clinical advisory group for deployment of the AI software.

Patients were not involved in the design of this study as it was conducted as a service evaluation to assess AI performance in prioritising chest X-rays. The evaluation focused on system-level implementation and did not directly impact patient care.

The statistical analysis assessed the prevalence of each of the AI findings in 2 patient cohorts. For the purposes of the analysis, the retrospective dataset is called the ‘lung cancer’ cohort, and the prospective dataset the ‘referral’ cohort. It is recognised these are separate patient cohorts and that the referral cohort will contain a small percentage of patients that will have a detectable lung cancer on CXR, estimated at < 1% based on Greater Manchester data and longitudinal cohort studies [11] [12].

- Lung cancer cohort: A retrospective dataset of all adult chest x-rays performed up to 6 months prior to a lung cancer diagnosis (4408 CXR, June 2020 to June 2022).
- Referral cohort: A consecutive prospective dataset of all chest x-rays performed in the adult population (107,065 CXR, March 2023 to June 2023).

The retrospective lung cancer cohort were obtained from the Greater Manchester cancer pathway registration dataset with ICD code 34.0. These data were matched with the diagnostic imaging data to identify all adult CXR performed within 6 months prior to the cancer diagnosis. The referral cohort is a consecutive dataset of all CXRs performed in Greater Manchester over a 3-month period from March 2023 to June 2023 which were also processed by the AI real-time at the time of CXR acquisition. We excluded CXRs for which AI results were not available in the extracted data. The datasets were further subdivided by the referral source to compare prevalence rates in patients presenting with CXR through General Practice (GP) and Accident & Emergency (A&E). The other CXR were from inpatient, day case and outpatient referral sources (Supplemental Figure 1). All data used in this study were anonymised prior to analysis to ensure patient privacy. Data processing adhered to GDPR regulations and NHS information governance standards to maintain confidentiality.

The main objective was to investigate the prevalence and the ratio of prevalence of AI findings in the lung cancer and referral cohorts. For comparing the prevalence of AI findings between the two cohorts, a two-sample chi-squared test for proportions was used. The prevalence ratio (PR) is defined as the ratio of prevalence of AI findings in lung cancer cohort to that of referral cohort. Both prevalences and prevalence ratios are reported. PR is analogous to the risk ratio (RR) of prospective cohort studies and case-control studies. The ‘Sensitivity’ of each AI finding for detecting cancer is the percentage of true positives in the cancer cohort.

### Matched Analysis

As the baseline characteristics may account for some of the difference in AI findings between the cohorts, we also conducted an adjusted analysis using propensity matched samples. A nearest neighbour matching method using scores from a multivariable logistic regression model fitted using age, gender, and referral source (A&E, GP or Other) as covariates was used for this. For each case in the lung cancer cohort, four matched cases from the referral cohort were selected (1:4 nearest neighbour matching). Thus, in addition to the prevalences and prevalence ratios in the overall sample, we also report them in the adjusted sample.

## Results

The baseline characteristics of age and gender show lung cancer cohort was an older age group 72 years mean ± 10 SD compared to the referral cohort 62 years ± 19 SD. There was no significant difference in the gender distribution of CXR in the cancer and referral cohorts, with a near equal distribution between the sexes. Analysis of CXR by referral source in the cancer cohort showed lung cancer presents most commonly following an A&E CXR referral, followed by GP CXR. The pre- defined AI priority groups show an expected distribution with Priority 1 and 2 findings being more prevalent in patients with lung cancer cohort than in the referral cohort (Table 2).

**Table 2:**
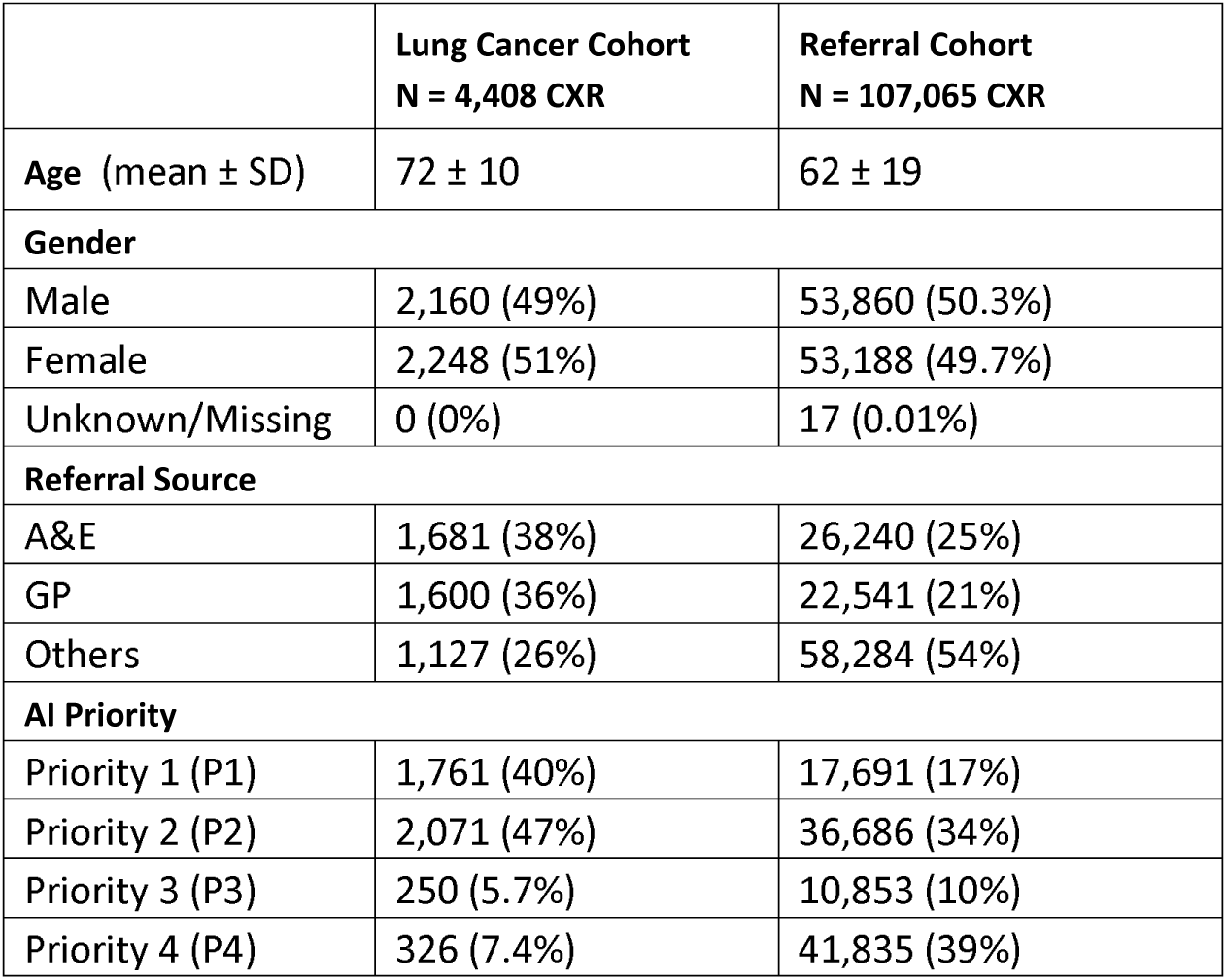
Baseline Characteristics.

**Table 3:**
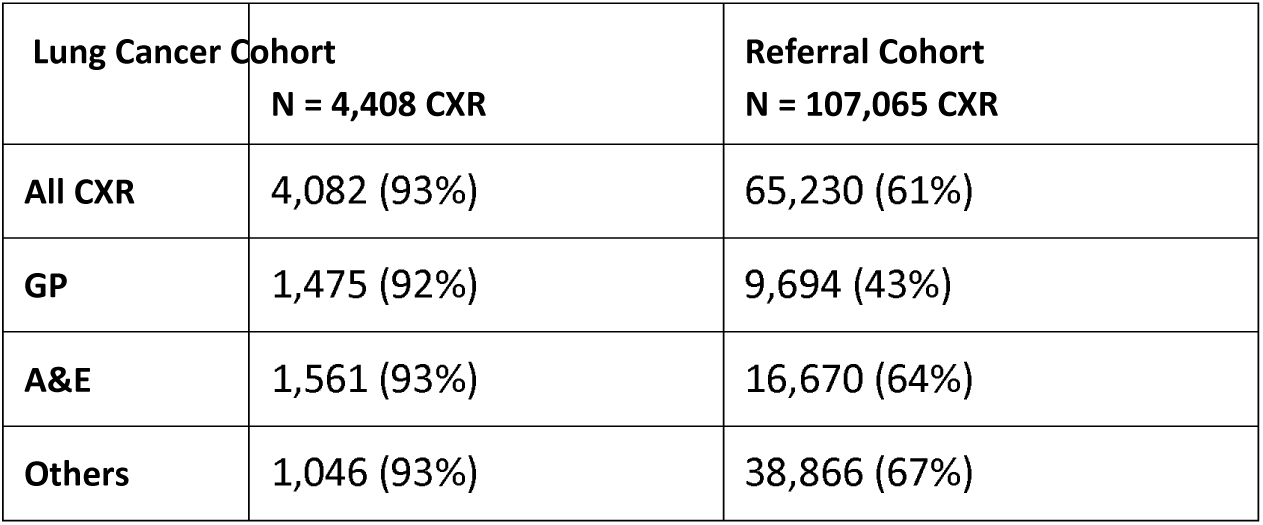
Number and percent of CXR classed as ‘Abnormal’ by AI.

|  | **Lung Cancer Cohort**<br>**N = 4,408 CXR** | **Referral Cohort**<br>**N = 107,065 CXR** |
| --- | --- | --- |
| **All CXR** | 4,082 (93%) | 65,230 (61%) |
| **GP** | 1,475 (92%) | 9,694 (43%) |
| **A&E** | 1,561 (93%) | 16,670 (64%) |
| **Others** | 1,046 (93%) | 38,866 (67%) |

The relative prevalence of AI priority findings in the lung cancer and referral cohorts gives an indication of the percentage of CXR that would need to be prioritised in the referral cohort to detect the corresponding percent of CXR in patients with cancer (Table 2).

- If CXR with AI Priority 1 were prioritised this would include 17% of CXR in the referral cohort and 40% of the CXR in the cancer cohort.
- Combining AI Priorities 1 and 2, prioritising 17 + 34 = 51% of CXR in the referral cohort would have included 40 + 47 = 87% of CXR in the cancer cohort.

CXR were classed as AI ‘Abnormal’ if the CXR has any of the detectable AI findings present. The relative number and percent of CXR classified as AI ‘Abnormal’ are higher in the A&E referral cohort (64%) compared to the GP referral cohort (43%). The difference in prevalence of AI ‘Abnormal’ between referral sources in the lung cancer cohort did not show any notable differences (Table3).

The prevalence of each individual AI finding was assessed in the lung cancer cohort and referral cohort. It is noted that no individual AI finding was present in more than half of the CXR in patients with lung cancer (Supplemental tables 1–3).

Combinations of AI findings produce a higher combined prevalence percentage in both the cancer cohort and referral cohort. This is not a direct summation of the prevalence of the individual AI findings as they often occur in combination.

All AI findings showed a statistical difference in prevalence (2-sample chi squared test for difference in proportions, p < 0.001) comparing the lung cancer and referral cohorts, except for cardiomegaly, mediastinal widening, pneumoperitoneum, and rib fracture (p > 0.05).

### Matched Sample Analysis Results

The propensity matched sample based on 1:4 nearest neighbour matching had all the 4408 CXRs from the lung cancer cohort and 17,632 matched CXRs from the referral cohort (Supplemental table 4). The absolute differences between the covariates (age, gender, and referral source) were more or less close to zero after the matching (Supplemental Figure 2).

The relative difference in prevalence of AI priority findings between the cancer and referral cohorts was less in the matched sample (Supplemental table 5). The prevalence of the individual AI findings in this matched adjusted sample are included in the Supplemental tables (Supplemental tables 6–8).

### Modelling combinations of AI findings to prioritise CXR for lung cancer

The initial group of AI findings to detect lung cancer was based on expert lung clinician and radiologist advice on the AI findings we would expect to be associated with significant pathologies in clinical practice.

The prevalence ratio (PR) correlates with each of the AI priorities for each referral group, suggesting the AI findings chosen by the expert clinicians are appropriate for the intended purpose (Table 4).

**Table 4:** Prevalence ratios – the relative percentage prevalence of CXR in each AI Priority group comparing the lung cancer patient cohort with the referral patient cohort.

| AI Priority | Lung Cancer (N=4408) | Referral Cohort (N=107065) | % in Lung Cancer | % in Referral Cohort | Prevalence Ratio (PR) (95% CI) |
| --- | --- | --- | --- | --- | --- |
| **All CXR** |  |  |  |  |  |
| P1 | 1761 | 17691 | 39.95 | 16.52 | 2.42 (2.33–2.51) |
| P2 | 2071 | 36686 | 46.98 | 34.27 | 1.37 (1.33-1.42) |
| P3 | 250 | 10853 | 5.67 | 10.14 | 0.56 (0.49-0.63) |
| P4 | 326 | 41835 | 7.40 | 39.07 | 0.19 (0.17-0.21) |
| **GP Referral CXR** |  |  |  |  |  |
| P1 | 766 | 2695 | 47.88 | 11.96 | 4.0 (3.76-4.26) |
| P2 | 633 | 4576 | 39.56 | 20.30 | 1.95 (1.82-2.08) |
| P3 | 76 | 2423 | 4.75 | 10.75 | 0.44 (0.25-0.55) |
| P4 | 125 | 12847 | 7.81 | 56.99 | 0.14 (0.12-0.16) |
| **A&E Referral CXRs** |  |  |  |  |  |
| P1 | 617 | 4479 | 36.70 | 17.07 | 2.15 (2.01-2.30) |
| P2 | 822 | 9194 | 48.90 | 35.04 | 1.39 (1.32-1.47) |
| P3 | 122 | 2997 | 7.26 | 11.42 | 0.63 (0.53-0.76) |
| P4 | 120 | 9570 | 7.14 | 36.47 | 0.20 (0.16-0.23) |

In clinical practice it is intended that both Priority 1 and Priority 2 cases are prioritised. The prevalence data indicate the following percent of CXR would need to be prioritised in each cohort. For example, for GP referral CXR, to include 86% of lung cancers in the cancer cohort we would need to prioritise 32% (P1 and P2) GP CXR in the referral cohort. This would require a lower number of patients to be prioritised compared with including all AI abnormal, 43% of CXR (Table 5).

**Table 5:** Prioritisation modelling showing the indicative percent of patient CXR that would be prioritised in the referral and lung cancer cohort for overall and stratified by referral sources of GP and A&E.

| CXR Referral Source | Total in Lung Cancer Cohort | Total in Referral Cohort | % P1 or P2 Lung Cancer Cohort | % P1 or P2 Referral Cohort | Prevalence Ratio (PR) (95% CI) |
| --- | --- | --- | --- | --- | --- |
| **Prioritising AI priority P1 and P2 CXR** |  |  |  |  |  |
| GP | 1600 | 22541 | 1399 (87%) | 7271 (32%) | 2.71<br>(2.64-2.78) |
| A&E | 1681 | 26240 | 1439 (86%) | 13673 (52%) | 1.64<br>(1.61-1.68) |
| Others | 1127 | 58284 | 994 (88%) | 33433 (57%) | 1.54<br>(1.50-1.57) |
| All CXR | 4408 | 107065 | 3882 (87%) | 54377 (51%) | 1.73<br>(1.71-1.75) |
| **Prioritising All Abnormal CXR** |  |  |  |  |  |
| GP | 1600 | 22541 | 1475 (92%) | 9694 (43%) | 2.14<br>(2.10-2.19) |
| A&E | 1681 | 26240 | 1561 (93%) | 16670 (64%) | 1.46<br>(1.44-1.48) |
| Others | 1127 | 58284 | 1046 (93%) | 38866 (67%) | 1.39<br>(1.37-1.42) |
| All CXR | 4408 | 107065 | 4082 (93%) | 65230 (61%) | 1.52<br>(1.50-1.53) |

We also modelled using a total of 106,473 distinct combinations (153 for two combinations, 816 for 3 combinations, 3060 for 4 combinations, 8568 for 5 combinations, 18564 for 6 combinations, 31824 for 7 combinations, 43758 for 8 combinations) of the 18 AI findings (excluding ‘Abnormal’ AI finding) to investigate the prevalence and prevalence ratio of various plausible combinations. The detailed findings of these are beyond the scope of the paper but on review the clinical advisory group considered there was equivocal benefit in adjusting the predefined categories based purely on cancer prevalence, as the groupings were set up to include AI findings needing priority review but not necessarily associated with lung cancer (e.g. pneumothorax and pneumoperitoneum).

It was agreed to elevate the AI finding ‘Consolidation’ to Priority 2, although this does not significantly change the numbers prioritised as nearly all CXR with consolidation also included the less specific AI finding of ‘Opacity’, which was already categorised as a Priority 2 finding. The AI finding ‘Tracheal deviation’ should also be prioritised, although the numbers are relatively small, and this has little impact on the overall percentage of patients requiring to be prioritised.

## Discussion

The NICE evaluation of Artificial intelligence-derived software to analyse chest X-rays for suspected lung cancer in primary care referrals has identified the need for more research to identifying normal or abnormal images, highlighting suspected abnormalities, prioritising review of CXR and speeding up subsequent referral to CT scan [10]. The ability of AI to detect abnormal lung features such as lung nodules, opacities, pleural effusion, collapsed lung segments, destruction, or erosion of bony structures such as the ribs, as well as masses or lymph nodes within the mediastinum could help prioritise abnormal CXR for rapid reporting.

Sites evaluating AI products need to be aware of the strengths and limitations of AI and plan their deployment and workflows to optimise the faster diagnosis of lung cancer, whilst minimising any detrimental effect on patients that are not prioritised. As no AI is 100% sensitive, any prioritisation based on the findings will benefit some patients over others in the CXR reporting queue [13].

This study provides a template methodology for how sites can determine the relative prevalence of AI findings in patients with cancer, compared to the prevalence of the AI findings in the referral population. This can help determine the local prioritisation strategy, inform ethical considerations around prioritisation, and indicate the capacity required to report the prioritised cases. The ability to monitor the prevalence of AI findings in a referral cohort is also important for post market surveillance, to identify potential performance drift with AI upgrades and potential changes in the referral cohort demographic [10].

Lung cancer can manifest in different ways on a CXR, and our observation that no individual AI finding was present in more than 50% of patient with lung cancer indicates that prioritisation strategies should include a combination of AI findings. Strategies should also consider other critical findings that may need to be prioritised, such as pneumoperitoneum or pneumothorax, that may not relate to the cancer diagnosis. In practice, we would wish to detect the maximum number of CXR with findings associated with cancer and other significant pathologies, while trying to keep the prevalence ratio high to avoid too many CXR being prioritised from the referral population.

This study has validated a potential Priority 1-4 model that could be considered. The prevalence and accuracy of individual AI findings will vary between AI software provider, and it is advised that a local assessment is performed in ‘shadow mode’ for each supplier. Vendor neutral AI platforms, such as proposed by the NHS AI lab, may also support collation of sample datasets in patients with cancer and enable benchmarking of AI performance between different AI software products [12].

The number of patients that may need to be prioritised to identify patients with lung cancer is significantly higher than the incidence of lung cancers in the referral population. This will vary depending on the referral source and findings to be prioritised. Adopting a model of prioritising patients with Priority 1 and 2 findings in this study would require 32% of GP referral CXR and 52% of A&E referral CXR to be prioritised respectively, to include the < 1% cancers detectable on CXR in the referral population. This reflects that each of the findings - nodule, consolidation etc. are not specific to cancer and may be found in other conditions such as infections and granulomatous diseases [15]. Modelling of CXR resource allocation and turnaround time was performed based on these data to inform our local prioritisation strategy but is outside the scope of this paper.

A limitation of this study is that the cancer and referral cohorts are from different times, the cancer cohort retrospective from 2020-22 and the referral cohort prospective from 2023. This enables the study to report only indicative prevalence ratios, and not predictive risk ratios reported in longitudinal prospective cohort studies that follow up patients to see who develops cancer. This approach was taken intentionally as it is a method that can be used by sites as an initial pre-clinical deployment ‘shadow mode’ analysis to inform the subsequent clinical deployment prioritisation. A longitudinal prospective study would take several years to identify the cancer cohort and would not be able to inform the local prioritisation strategy and model at the outset.

We conclude that AI can identify findings associated with lung cancer at the time of chest x-ray acquisition and prioritise them for rapid reporting. The number of cases requiring prioritisation depends on the prevalence of AI findings in the referral population and the prevalence in patients with the disease. This paper sets out the methodology and a sample prioritisation model that can be used to inform and validate which AI finding should be prioritised.

### Author contributions

RB was the clinical project lead and is a consultant radiologist and diagnostics, digital and innovation lead with the Greater Manchester (GM) Cancer Alliance. AS is a lead lung cancer radiologist in GM. PB is a radiology registrar in GM. RN, SG, LB ME are consultant lung physicians were working with the GM Cancer Alliance and lead members of the lung cancer pathway board. EW provided technical and analytical project support. DR provided the statistical analyses. MS provided the sponsor funding support and oversight. RB led the write up of the manuscript and all authors agreed the final version.

## Data Availability

All data produced in the present work are contained in the manuscript and supplemental tables

## Acknowledgements

The authors would like to acknowledge the support of the GM Cancer Alliance, project sponsor and suppliers that worked collaboratively throughout the project. Sarah Lyon was the GM cancer alliance project manager and Lisa Galligan Dawson the GM cancer performance director and senior responsible officer. Rohit Agrawal and Surabhi provided project support within Qure.ai. Nynke Breimer provided PACS platform support as the global product manager for the Sectra Amplifier Services.

## Supplemental Tables

### Overall sample

### Propensity matched sample

**Supplemental Figure 1:**
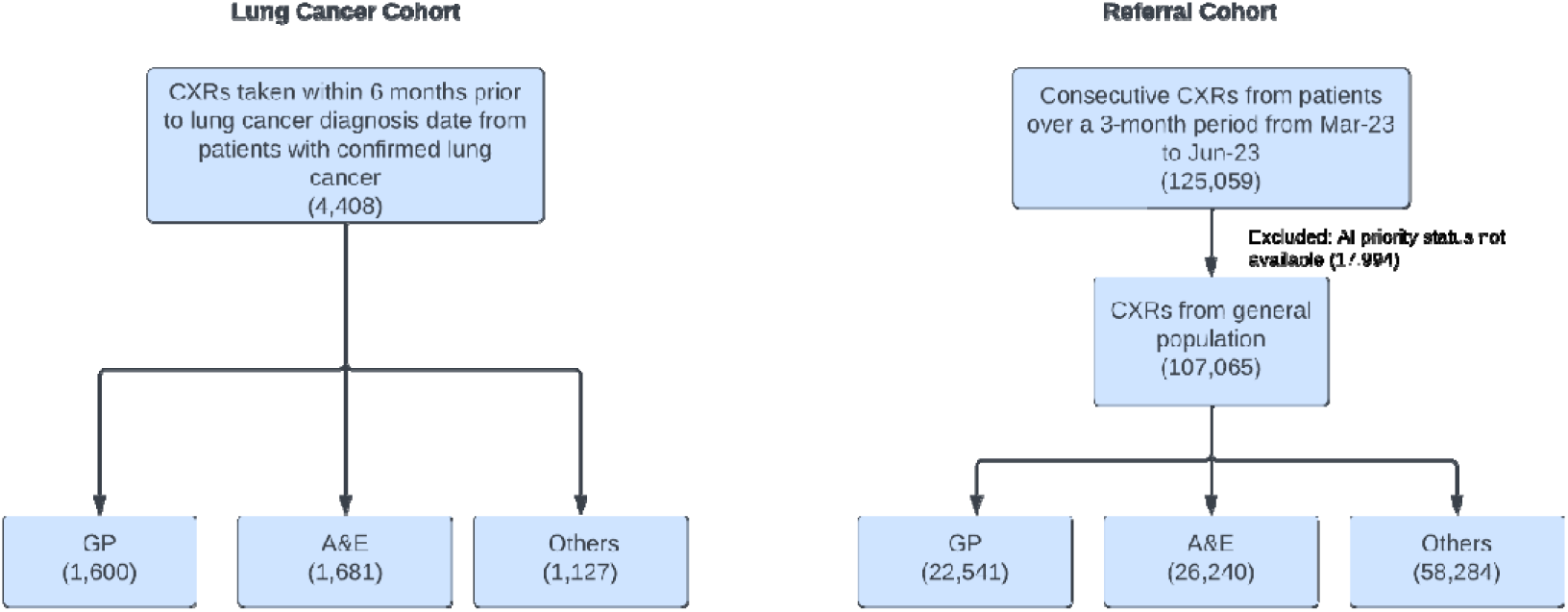
Derivation diagram of the CXR for the retrospective lung cancer patient cohort and prospective referral cohort from the general population, subdivided by referral source.

**Supplemental Figure 2:**
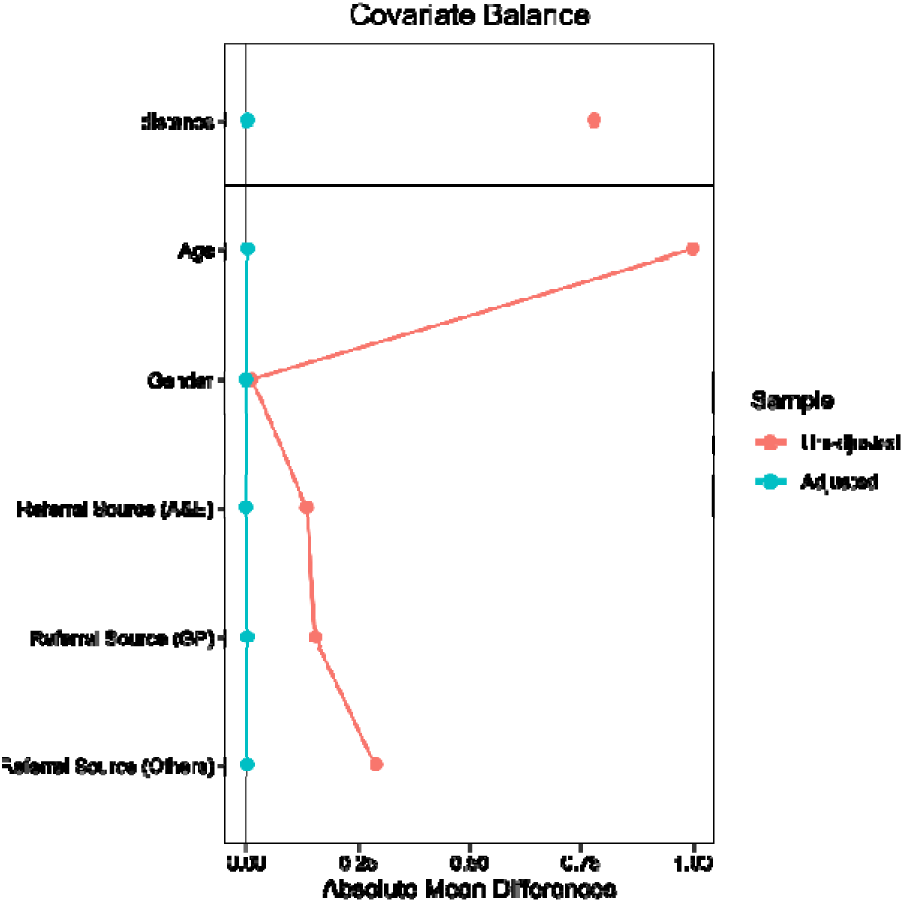
Covariate balance love plot for patient age, gender and referral source comparing the unadjusted and propensity matched adjusted referral patient cohort.

**Supplemental Table 1:** Proportion of AI Findings (All referral sources)

|  | Lung Cancer (N) | Referral Cohort (N) | % in Lung Cancer | % in Referral Cohort | p* |
| --- | --- | --- | --- | --- | --- |
| AI Priority 1 | 1761 | 17691 | 39.95 | 16.52 | <0.00001 |
| AI Priority 2 | 2071 | 36686 | 46.98 | 34.27 | <0.00001 |
| AI Priority 3 | 250 | 10853 | 5.67 | 10.14 | <0.00001 |
| AI Priority 4 | 326 | 41835 | 7.40 | 39.07 | <0.00001 |
| Abnormal | 4082 | 65230 | 92.60 | 60.93 | <0.00001 |
| Opacity | 3678 | 48549 | 83.44 | 45.35 | <0.00001 |
| Nodule | 1439 | 11707 | 32.65 | 10.93 | <0.00001 |
| Lung Nodule Malignancy | 522 | 956 | 11.84 | 0.89 | <0.00001 |
| Cardiomegaly | 1070 | 27334 | 24.27 | 25.53 | >0.05 |
| Pneumoperitoneum | 46 | 1147 | 1.04 | 1.07 | >0.05 |
| Emphysema | 1182 | 11650 | 26.81 | 10.88 | <0.00001 |
| Reticulonodular Pattern | 575 | 828 | 13.04 | 8.25 | <0.00001 |
| Atelectasis | 1476 | 22352 | 33.48 | 20.88 | <0.00001 |
| Mediastinal Widening | 242 | 5382 | 5.49 | 5.03 | >0.05 |
| Rib Fracture | 247 | 5590 | 5.60 | 5.22 | >0.05 |
| Hilar Enlargement | 94 | 1154 | 2.13 | 1.08 | <0.00001 |
| Pleural Effusion | 1246 | 23639 | 28.27 | 22.08 | <0.00001 |
| Fibrosis | 1255 | 11571 | 28.47 | 10.81 | <0.00001 |
| Blunted Costophrenic Angle | 130 | 2254 | 2.95 | 2.11 | <0.001 |
| Pneumothorax | 214 | 4115 | 4.85 | 3.84 | <0.001 |
| Cavity | 112 | 675 | 2.54 | 0.63 | <0.00001 |
| Consolidation | 1626 | 13795 | 36.89 | 12.88 | <0.00001 |
| Tracheal Deviation | 261 | 2032 | 5.92 | 1.90 | <0.00001 |

\*p-value based on the 2-sample chisquare test for testing the difference in proportions in lung cancer and referral cohort.

**Supplemental Table 2:** Proportion of AI findings (A&E CXR referrals)

|  | Lung Cancer (N) | Referral Cohort (N) | % in Lung Cancer | % in Referral Cohort | P* |
| --- | --- | --- | --- | --- | --- |
| Priority 1 | 617 | 4479 | 36.70 | 17.07 | <0.00001 |
| Priority 2 | 822 | 9194 | 48.90 | 35.04 | <0.00001 |
| Priority 3 | 122 | 2997 | 7.26 | 11.42 | <0.00001 |
| Priority 4 | 120 | 9570 | 7.14 | 36.47 | <0.00001 |
| Abnormal | 1561 | 16670 | 92.86 | 63.53 | <0.00001 |
| Opacity | 1371 | 12153 | 81.56 | 46.31 | <0.00001 |
| Nodule | 464 | 2722 | 27.60 | 10.37 | <0.00001 |
| Lung Nodule Malignancy | 151 | 218 | 8.98 | 0.83 | <0.00001 |
| Cardiomegaly | 540 | 8014 | 32.12 | 30.54 | >0.05 |
| Pneumoperitoneum | 16 | 259 | 0.95 | 0.99 | >0.05 |
| Emphysema | 440 | 2863 | 26.17 | 10.91 | <0.00001 |
| Reticulonodular Pattern | 253 | 2308 | 15.05 | 8.80 | <0.00001 |
| Atelectasis | 574 | 5678 | 34.15 | 21.64 | <0.00001 |
| Mediastinal Widening | 124 | 1532 | 7.38 | 5.84 | <0.05 |
| Rib Fracture | 118 | 1571 | 7.02 | 5.99 | >0.05 |
| Hilar Enlargement | 40 | 433 | 2.38 | 1.65 | <0.05 |
| Pleural Effusion | 508 | 5606 | 30.22 | 21.36 | <0.00001 |
| Fibrosis | 432 | 2537 | 25.70 | 9.67 | <0.00001 |
| Blunted Costophrenic Angle | 37 | 459 | 2.20 | 1.75 | >0.05 |
| Pneumothorax | 56 | 799 | 3.33 | 3.04 | >0.05 |
| Cavity | 38 | 136 | 2.26 | 0.52 | <0.00001 |
| Consolidation | 564 | 2982 | 33.55 | 11.36 | <0.00001 |
| Tracheal Deviation | 104 | 505 | 6.19 | 1.92 | <0.00001 |

\*p-value based on the 2-sample chisquare test for testing the difference in proportions in lung cancer and referral cohort.

**Supplemental Table 3:** Proportion of Findings (GP CXR referrals)

|  | Lung Cancer (N) | Referral Cohort (N) | % in Lung Cancer | % in Referral Cohort | p* |
| --- | --- | --- | --- | --- | --- |
| Priority 1 | 766 | 2695 | 47.88 | 11.96 | <0.00001 |
| Priority 2 | 633 | 4576 | 39.56 | 20.30 | <0.00001 |
| Priority 3 | 76 | 2423 | 4.75 | 10.75 | <0.00001 |
| Priority 4 | 125 | 12847 | 7.81 | 56.99 | <0.00001 |
| Abnormal | 1475 | 9694 | 92.19 | 43.01 | <0.00001 |
| Opacity | 1347 | 6322 | 84.19 | 28.05 | <0.00001 |
| Nodule | 675 | 2302 | 42.19 | 10.21 | <0.00001 |
| Lung Nodule Malignancy | 278 | 131 | 17.38 | 0.58 | <0.00001 |
| Cardiomegaly | 215 | 2379 | 13.44 | 10.55 | <0.001 |
| Pneumoperitoneum | 11 | 78 | 0.69 | 0.35 | <0.05 |
| Emphysema | 483 | 2657 | 30.19 | 11.79 | <0.00001 |
| Reticulonodular Pattern | 154 | 755 | 9.62 | 3.35 | <0.00001 |
| Atelectasis | 510 | 2734 | 31.87 | 12.13 | <0.00001 |
| Mediastinal Widening | 50 | 321 | 3.12 | 1.42 | <0.00001 |
| Rib Fracture | 58 | 552 | 3.62 | 2.45 | <0.01 |
| Hilar Enlargement | 37 | 86 | 2.31 | 0.38 | <0.00001 |
| Pleural Effusion | 343 | 1573 | 21.44 | 6.98 | <0.00001 |
| Fibrosis | 511 | 1974 | 31.94 | 8.76 | <0.00001 |
| Blunted Costophrenic Angle | 55 | 488 | 3.44 | 2.16 | <0.01 |
| Pneumothorax | 34 | 210 | 2.12 | 0.93 | <0.00001 |
| Cavity | 53 | 78 | 3.31 | 0.35 | <0.00001 |
| Consolidation | 644 | 690 | 40.25 | 3.06 | <0.00001 |
| Tracheal Deviation | 88 | 140 | 5.50 | 0.62 | <0.00001 |

\*p-value based on the 2-sample chisquare test for testing the difference in proportions in lung cancer and referral cohort.

**Supplemental Table 4:** Number of patients in the overall and the 1:4 propensity matched cohort.

|  | Lung Cancer | Referral Cohort |
| --- | --- | --- |
| **All** | 4408 | 107065 |
| **Matched** | 4408 | 17632 |
| **Unmatched** | 0 | 89416 |
| **Discarded** | 0 | 17* |

\*17 CXRs were excluded from the original 107,065 referral cohort due to missing age.

**Supplemental Table 5:** Proportion of AI Findings in Unadjusted and Matched Sample.

| Priority Status | UNADJUSTED SAMPLE |  |  | MATCHED SAMPLE |  |  |
| --- | --- | --- | --- | --- | --- | --- |
|  | Lung Cancer Cohort (%) | Referral Cohort (%) | p* | Lung Cancer Cohort (%) | Referral Cohort (%) | p* |
| **P1** | 39.95 | 16.52 | <0.00001 | 39.95 | 19.30 | <0.00001 |
| **P2** | 46.98 | 34.27 | <0.00001 | 46.98 | 37.68 | <0.00001 |
| **P3** | 5.67 | 10.14 | <0.00001 | 5.67 | 12.26 | <0.00001 |
| **P4** | 7.40 | 39.07 | <0.00001 | 7.40 | 30.76 | <0.00001 |

\*p-value based on the two-sample chisquare test for testing the difference in proportions in lung cancer and referral cohort.

**Supplemental Table 6:**
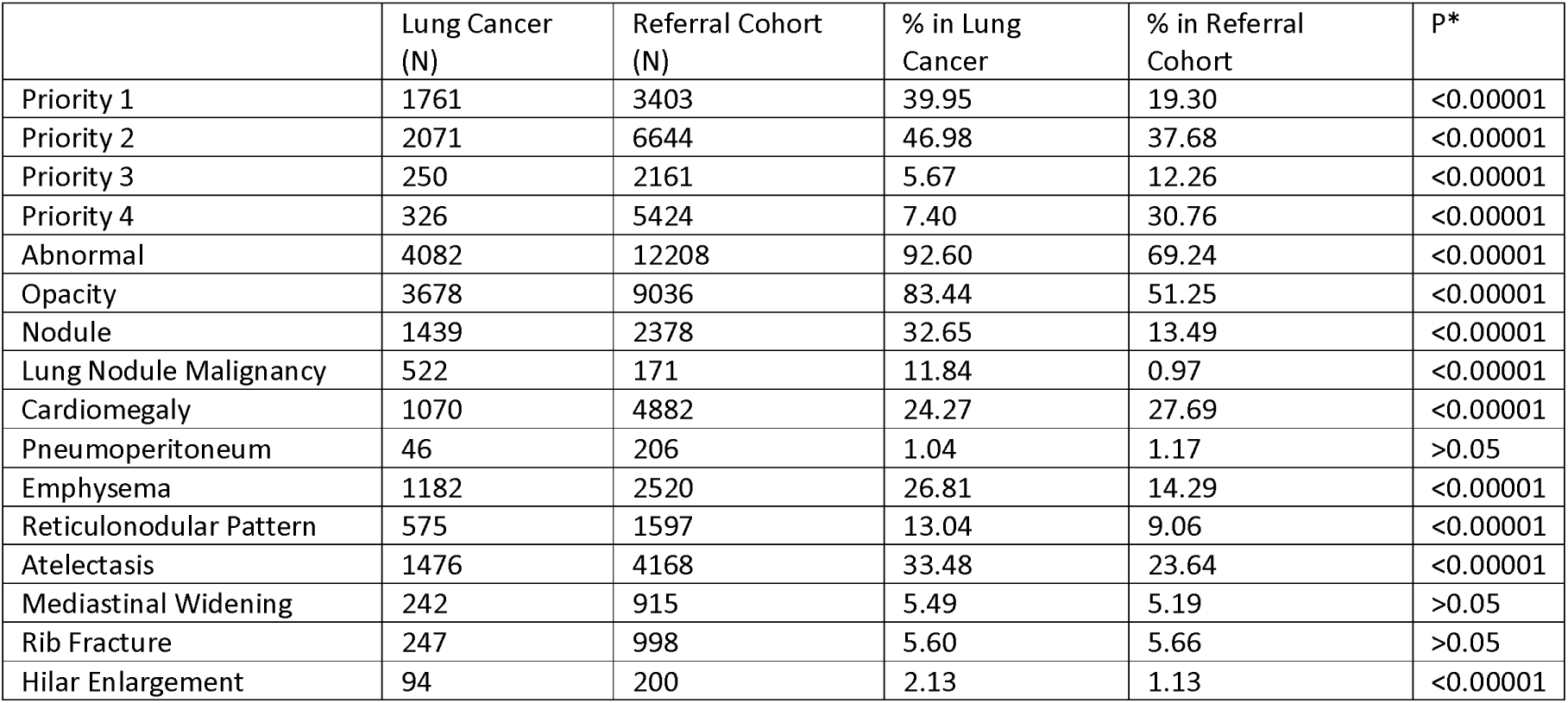
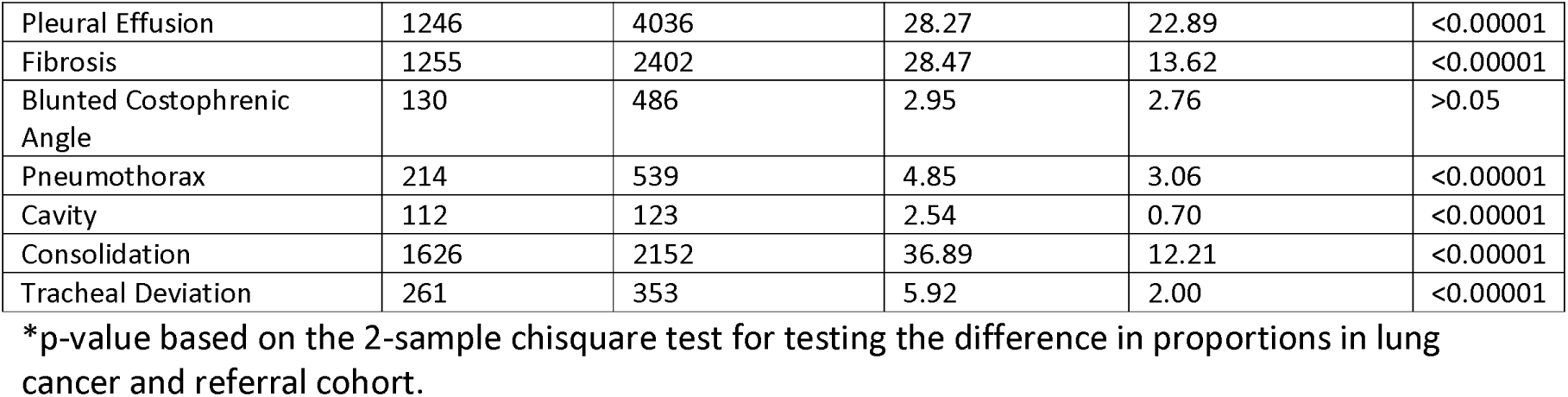
Proportion of AI findings (propensity matched sample – all patients)

**Supplemental Table7:** Proportion of AI findings (propensity matched sample – A&E patients)

|  | Lung Cancer (N) | Referral Cohort (N) | % in Lung Cancer | % in Referral Cohort | P* |
| --- | --- | --- | --- | --- | --- |
| Priority 1 | 617 | 1405 | 36.70 | 20.89 | <0.00001 |
| Priority 2 | 822 | 2864 | 48.90 | 42.59 | <0.00001 |
| Priority 3 | 122 | 880 | 7.26 | 13.09 | <0.00001 |
| Priority 4 | 120 | 1576 | 7.14 | 23.43 | <0.00001 |
| Abnormal | 1561 | 5149 | 92.86 | 76.57 | <0.00001 |
| Opacity | 1371 | 3818 | 81.56 | 56.77 | <0.00001 |
| Nodule | 464 | 857 | 27.60 | 12.74 | <0.00001 |
| Lung Nodule Malignancy | 151 | 64 | 8.98 | 0.95 | <0.00001 |
| Cardiomegaly | 540 | 2532 | 32.12 | 37.65 | <0.001 |
| Pneumoperitoneum | 16 | 81 | 0.95 | 1.20 | >0.05 |
| Emphysema | 440 | 943 | 26.17 | 14.02 | <0.00001 |
| Reticulonodular Pattern | 253 | 749 | 15.05 | 11.14 | <0.001 |
| Atelectasis | 574 | 1813 | 34.15 | 26.96 | <0.00001 |
| Mediastinal Widening | 124 | 484 | 7.38 | 7.20 | >0.05 |
| Rib Fracture | 118 | 458 | 7.02 | 6.81 | >0.05 |
| Hilar Enlargement | 40 | 125 | 2.38 | 1.86 | >0.05 |
| Pleural Effusion | 508 | 1822 | 30.22 | 27.09 | <0.05 |
| Fibrosis | 432 | 879 | 25.70 | 13.07 | <0.00001 |
| Blunted Costophrenic Angle | 37 | 147 | 2.20 | 2.19 | >0.05 |
| Pneumothorax | 56 | 236 | 3.33 | 3.51 | >0.05 |
| Cavity | 38 | 41 | 2.26 | 0.61 | <0.00001 |
| Consolidation | 564 | 959 | 33.55 | 14.26 | <0.00001 |
| Tracheal Deviation | 104 | 162 | 6.19 | 2.41 | <0.00001 |

\*p-value based on the 2-sample chisquare test for testing the difference in proportions in lung cancer and referral cohort.

**Supplemental Table 8:** Proportion of AI findings (propensity matched sample – GP patients)

|  | Lung Cancer (N) | Referral Cohort (N) | % in Lung Cancer | % in Referral Cohort | P* |
| --- | --- | --- | --- | --- | --- |
| Priority 1 | 766 | 1018 | 47.88 | 15.91 | <0.00001 |
| Priority 2 | 633 | 1744 | 39.56 | 27.25 | <0.00001 |
| Priority 3 | 76 | 861 | 4.75 | 13.46 | <0.00001 |
| Priority 4 | 125 | 2776 | 7.81 | 43.38 | <0.00001 |
| Abnormal | 1475 | 3623 | 92.19 | 56.62 | <0.00001 |
| Opacity | 1347 | 2452 | 84.19 | 38.32 | <0.00001 |
| Nodule | 675 | 883 | 42.19 | 13.80 | <0.00001 |
| Lung Nodule Malignancy | 278 | 45 | 17.38 | 0.70 | <0.00001 |
| Cardiomegaly | 215 | 916 | 13.44 | 14.31 | >0.05 |
| Pneumoperitoneum | 11 | 46 | 0.69 | 0.72 | >0.05 |
| Emphysema | 483 | 1028 | 30.19 | 16.07 | <0.00001 |
| Reticulonodular Pattern | 154 | 331 | 9.62 | 5.17 | <0.00001 |
| Atelectasis | 510 | 1109 | 31.87 | 17.33 | <0.00001 |
| Mediastinal Widening | 50 | 125 | 3.12 | 1.95 | <0.01 |
| Rib Fracture | 58 | 224 | 3.62 | 3.50 | >0.05 |
| Hilar Enlargement | 37 | 22 | 2.31 | 0.34 | <0.00001 |
| Pleural Effusion | 343 | 667 | 21.44 | 10.42 | <0.00001 |
| Fibrosis | 511 | 787 | 31.94 | 12.30 | <0.00001 |
| Blunted Costophrenic Angle | 55 | 186 | 3.44 | 2.91 | >0.05 |
| Pneumothorax | 34 | 72 | 2.12 | 1.13 | <0.01 |
| Cavity | 53 | 27 | 3.31 | 0.42 | <0.00001 |
| Consolidation | 644 | 269 | 40.25 | 4.20 | <0.00001 |
| Tracheal Deviation | 88 | 52 | 5.50 | 0.81 | <0.00001 |

\*p-value based on the 2-sample chisquare test for testing the difference in proportions in lung cancer and referral cohort.

## References

[1] CRUK, “Lung Cancer Statistics,” 13 March 2024. [Online]. Available: https://www.cancerresearchuk.org/health-professional/cancer-statistics/statistics-by-cancer-type/lung-cancer.

[2] NICE, “Suspected cancer: recognition and referral,” 2 October 2023. [Online]. Available: https://www.nice.org.uk/guidance/ng12.

[3] NHSE, “Implementing a timed lung cancer diagnostic pathway,” 12 October 2023. [Online]. Available: https://www.england.nhs.uk/long-read/implementing-a-timed-lung-cancer-diagnostic-pathway/.

[4] NHSE, “National Optimal Lung Cancer Pathway for suspected and confirmed lung cancer: Referral to treatment,” 2020. [Online]. Available: https://www.cancerresearchuk.org/sites/default/files/national_optimal_lung_pathway_aug_2017.pdf.

[5] NHSE, “Diagnostic Imaging Dataset Statistical Release [Feb 2024],” 22 February 2024. [Online]. Available: https://www.england.nhs.uk/statistics/wp-content/uploads/sites/2/2024/02/Statistical-Release-22nd-February-2024-PDF-305KB.pdf.

[6] A. Al, S. Zigman and F. Walter, “Experiences along the diagnostic pathway for patients with advanced lung cancer in the USA: a qualitative study,” BMJ Open, p. 11:e045056, 2021.

[7] N. Navani and D. Lawrence, “Lung cancer diagnosis and staging with endobronchial ultrasound-guided transbronchial needle aspiration compared with conventional approaches: an open-label, pragmatic, randomised controlled trial,” Lancet Respir Med, p. 3:282–9, 2015.

[8] M. Evison, K. Hewitt, J. Lyons, P. Crosbie, H. Balata, C. Gee, R. Duerden, M. Greaves, A. Sharman and R. Booton, “Implementation and outcomes of the RAPID programme: Addressing the front end of the lung cancer pathway in Manchester,” Clinical Medicine, pp. 20 (4) 401–405, Jul 2020.

[9] N. Woznitza, B. Ghimire and A. Devaraj, “Impact of radiographer immediate reporting of X-rays of the chest from general practice on the lung cancer pathway (radioX): a randomised controlled trial,” Thorax, pp. 78:890–894., 2023.

10. [10] NICE, “Artificial intelligence-derived software to analyse chest X-rays for suspected lung cancer in primary care referrals: early value assessment,” 28 September 2023. [Online]. Available: https://www.nice.org.uk/guidance/hte12.

[11] I. Baltruschat, L. Steinmaster and H. Nickisch, “Smart chest X-ray worklist prioritization using artificial intelligence: a clinical workflow simulation,” Eur Radiol, pp. Jun;31(6):3837–3845, 2021.

12. [12] NHSE, “The NHS AI Lab,” NHS England Transformation Directorate, [Online]. Available: https://transform.england.nhs.uk/ai-lab/. [Accessed 14 March 2024].

13. [13] M. Callister, Baldwin DR and A. Akram, “British Thoracic Society guidelines for the investigation and management of pulmonary nodules: accredited by NICE,” Thorax, pp. 70:ii1- ii54., 2015.

[14] S. Bradley, B. Bhartia and M. Callister, “Chest X-ray sensitivity and lung cancer outcomes: a retrospective observational study,” British Journal of General Practice, pp. 71 (712): e862–e868, 2021.

[15] S. Bradley, N. Hatton and B. Bhartia, “Estimating lung cancer risk from chest X-ray and symptoms: a prospective cohort study.,” Br J Gen Pract., pp. Mar 26;71(705):e280–e286, 2021.

